# Combination low dose sulphonylurea and DPP4 inhibitor have potent glucose lowering effect through augmentation of beta cell function without increase in hypoglycaemia: a randomised crossover study

**DOI:** 10.1101/2023.08.22.23294403

**Authors:** RLM Cordiner, K Bedair, A Mari, ER Pearson

**Affiliations:** Division of Population, Health and Genomics, Ninewells Hospital and Medical School, University of Dundee, United Kingdom; Institute of Neuroscience, National Research Council, University of Padua, Italy

**Author notes:** Corresponding Author Professor Ewan Pearson, Professor of Diabetic Medicine, Head of Division, Division of Population, Health, and Genomics, Ninewells Hospital and Medical School, University of Dundee, DD1 9SY. Tweet: Cordiner et al establish that combination of low dose sulphonylurea (20mg Gliclazide) and DPP4i have potent glucose lowering effect through augmentation of beta-cell function without increase in hypoglycaemia. @CordinerRuth @ezpearson @UoDMedicine.

## Abstract

**Aims/Hypothesis:** It is important to address our use of cheaper generic therapies as the global prevalence of type 2 diabetes (T2DM) will surpass 600 million by 2035. Negative aspects of SU may be avoided by their use at low dose. We have previously shown that 20mg standard release gliclazide reduces plasma glucose through augmentation of the classical incretin effect, increased beta-cell glucose sensitivity and late-phase incretin potentiation. We hypothesised that there would be potential synergy between low dose SU when given in combination with a DPP4i, without increased hypoglycaemia risk, and aimed to assess this in a randomised clinical trial.

**Methods:** 30 participants with T2DM (HbA1c <64 mmol/mol) treated with diet or metformin monotherapy were recruited to a single-centre, open-label, randomised crossover study. Participants completed four, 14-day study periods in a random order: control, gliclazide 20mg once daily (SU), sitagliptin 100mg (DPP4i), or combination (SUDPP4i). A 2-hour mixed meal tolerance test was conducted at the end of each block. Beta-cell function was assessed by modelling. The primary outcome was the effect of treatment on beta-cell glucose sensitivity. Secondary end points included frequency of blood glucose <3mmol/l on continuous glucose monitoring, sub analysis by genotype (KNCJ11 E23K), and analysis by gender and body mass index.

**Results:** Linear mixed model estimates showed a potent additive, glucose lowering effect of low dose SU combination with DPP4. Mean glucose AUC (mean 95% CI) (mmol/l) was: Control 11.5 (10.7 – 12.3), DPP4i 10.2 (9.4 – 11.1), SU 9.7 (8.9 – 10.5), SUDPP4i 8.7 (7.9 – 9.5) (p <0.001). Beta-cell glucose sensitivity (pmol min^-1^ m^-2^ mM^-1^) mirrored this additive effect: Control 71.5 (51.1 – 91.9), DPP4i 75.9 (55.7 – 96.0), SU 86.3 (66.1 – 106.4), SUDPP4i 94.1 (73.9 – 114.3) (p = 0.04). Glucose time in range <3mmol/l on CGM (%) was unaffected: Control 1 (2-4), DPP4i 2 (3-6), SU 1 (0-4), SUDPP4i 3 (2 – 7) (p = 0.65). The increase in glucose sensitivity with sulphonylurea treatment was seen in men not women.

**Conclusions:** Combination low dose gliclazide with a DPP4i has potent glucose lowering effect through augmentation of beta cell function. Glucose reduction was achieved at gliclazide concentrations far below those achieved with standard therapeutic doses. A double-blind randomised controlled trial is merited to formalise efficacy and safety of this combination, which may avoid negative aspects of SU and provide pharmacoeconomic benefit in diabetes care.

**Research in Context:** *What is already known about this subject?:* Previous isoglycaemic clamp studies in low dose sulphonylureas established that 20mg of gliclazide augments the classical incretin effect, increases glucose sensitivity by 50% and late phase incretin potentiation.

*What is the key question?:* What is the effect of low dose sulphonylureas as monotherapy or in combination with a DPP4i on parameters of beta cell function following a mixed meal?

*What are the new findings?:* Low dose sulphonylureas have potent glucose lowering potential which is further enhanced by the addition of a DPP4i, without increasing hypoglycaemia. Modelling of beta cell function demonstrates that low dose sulphonylureas heighten the beta cell dose response which is further augmented by the presence of a DPP4i. Phenotypic differences in response are noted, with male participants showing additional effect of glucose sensitivity in response to sulphonylureas. This effect is not seen in women. Gliclazide standard release at 20mg produces a similar pharmacokinetic profile during mixed meal tolerance test to 30mg of modified release gliclazide.

*How might this impact on clinical practice in the foreseeable future?:* These results suggest that it is possible to modernise the use of two cheap, effective second-line treatments of type 2 diabetes mellitus through future production of a combined preparation of low dose gliclazide and a DPP4i. This combination has real potential as a safe, efficacious treatment which could bring pharmacoeconomic benefit to low- and middle-income countries worldwide.

## Introduction (Current 4003)

Sulphonylureas (SU) have been utilised in the treatment of type 2 diabetes mellitus (T2DM) for over 70 years (1). However, their use has declined due to their associations with hypoglycaemia, weight gain, limited durability, and their lack of positive cardiovascular outcome data in comparison with newer agents. Currently, international guidelines recommend the use of SU “if cost is an issue” (2). However, the cost of diabetes care is escalating; the global prevalence of T2DM is predicted to increase from 382 million people to 592 million by 2035, which includes 69% increase in prevalence in developing countries and a 20% increase in developed countries (3, 4). The predicted absolute global economic burden of diabetes care will increase from $1.3 trillion US Dollars (95% confidence interval 1.3 – 1.3) in 2015 to $2.2 trillion (2.2 – 2.3) in 2030, which translates to an increase in costs as a share of the global GDP from 1.8% (1.7 – 1.9) in 2015 to a maximum of 2.2% (2.1 – 2.2) (5). This increase in per capita cost therefore poses a global emergency to control cost. A systematic review indicated that the economic burden of diabetes most directly affects patients in low- and middle-income countries (LMIC), with the magnitude of cost differing considerably between countries (6). Therefore, there is pharmacoeconomic need to provide cost-effective diabetes care, including how we modernise our use of our cheaper generic therapies such as SU and DPP4 inhibitors (DPP4i).

Studies in neonatal diabetes mellitus (NDM) have provided insight into the beta-cell response of SU. Studies in patients with NDM due to activating mutations in KCNJ11, found that these patients were able to effectively switch from insulin to high dose SU, with resulting tight glycaemic control whilst avoiding hypoglycaemia. Patients with NDM have no insulin response to intravenous glucose but have a significantly increased insulin response to oral glucose or mixed meal stimulus following SU initiation, suggesting augmentation of the incretin effect (7, 8). In addition, a supra-additive effect of SU in combination with high concentrations of intravenous GIP has also been shown in HNF1A-maturity onset diabetes of the young (HNF1A MODY) (9) and in T2DM treated with standard dose glipizide (10). Our previous work has shown that low dose SU augment the incretin effect (11). Using isoglycaemic clamps in patients with T2DM treated with diet or metformin monotherapy, we demonstrated that a 20mg dose of gliclazide reduced mean glucose AUC during oral glucose tolerance test from 12.0 to 10.8 mmol/l (*p*=0.0006), augmented the incretin effect from 35.5 to 55% (*p*=0.04), and increased glucose sensitivity by 50% (*p*=0.01) and enhanced late phase incretin potentiation (*p*=0.04).

Given that we have uncovered a novel mechanism of SU at low dose that results in glucose regulated insulin secretion, in part mediated by the incretin effect, we hypothesised that DPP4i, which increase endogenous incretins, would be a potent drug to combine with low dose SU. We aimed to explore the efficacy of low dose SU and endogenous incretins, as monotherapy, and in combination with a DPP4i on parameters of beta cell function utilising multiple mixed meal tolerance tests (MMT), beta cell modelling and continuous glucose monitoring (CGM).

## Methods

### Recruitment

30 participants were recruited with physician diagnosed T2DM treated with diet or metformin monotherapy, HbA1c <64 mmol/mol, aged ≥ 40 and ≤ 80 years and with renal and hepatic function in the biochemical reference range from local laboratories. To avoid heterogeneity within the cohort, only White British participants were recruited. All participants had capacity to express informed, written consent. Participants not meeting inclusion criteria were excluded. Patients who were pregnant, lactating or planning to conceive within the study period were ineligible. Patients participating or who were recruited in a clinical study within the preceding 30 days were also ineligible.

### Study Design

We undertook a single-site, open label, randomised crossover study involving MMT (Figure 1). The study was approved by The East of Scotland Research Ethics Committee (REC 18/ES/0092) and registered with ClinicalTrials.gov (NCT04192292). All research was conducted in accordance with the Declaration of Helsinki, and informed written consent was obtained for all participants prior to study inclusion.

**Figure 1.**
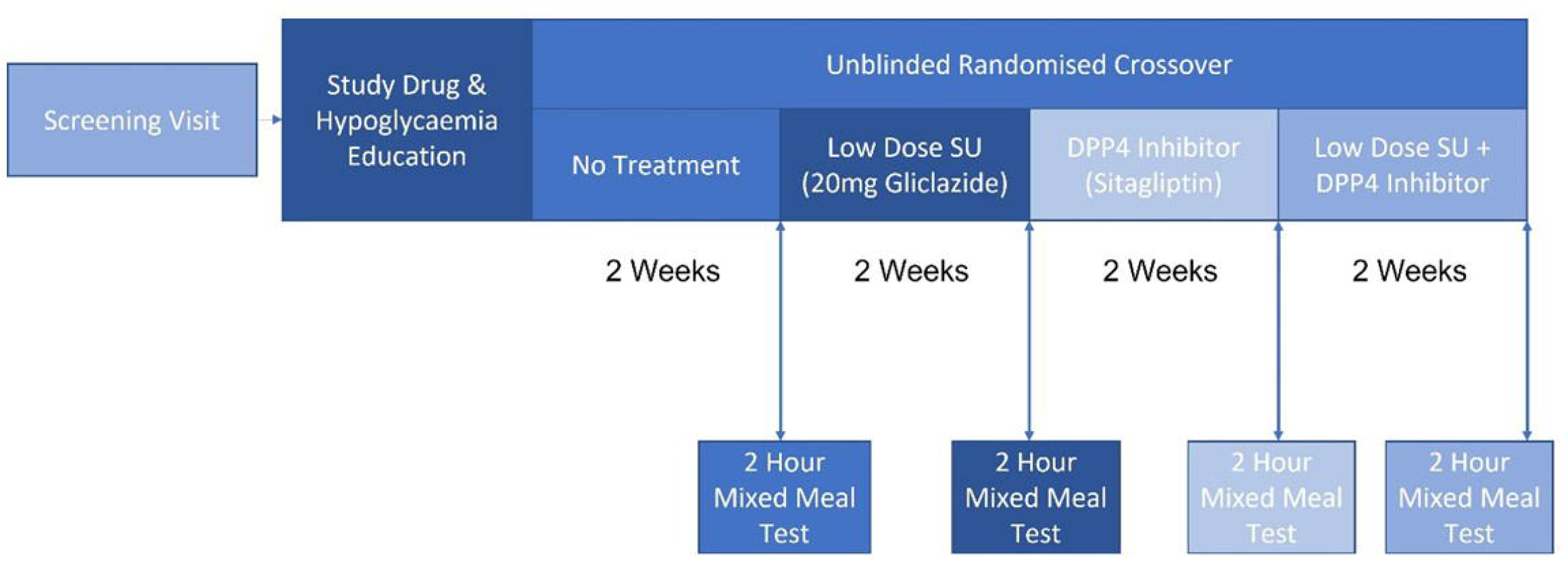
Unblinded randomised crossover study design involving four different study intervention periods, each 14 days duration. Participants completed a 2-hour mixed meal tolerance test at the end of each study period.

Study visits took place at The Clinical Research Centre, Ninewells Hospital and Medical School. The study involved four intervention blocks, each of 14-days duration, to assess response following different combination of low dose SU or DPP4i: no intervention (no change to standard care), low dose SU (20mg gliclazide once daily), DPP4i (100mg sitagliptin once daily), low dose SU + DPP4i (20mg gliclazide + 100mg sitagliptin once daily).

Participants attended the research centre on 6 separate visits. A screening visits confirmed eligibility and obtained informed written consent. The second visit provided education regarding study drugs, self-monitoring of blood glucose, and hypoglycaemia. The other four visits were performed at the end of each block.

Participants underwent a 2-hour MMT at the end of each block. Participants were fasted for 8-hours prior to intervention and all regular medications, including metformin, were withheld until the end of the test. On arrival to the centre, a single intravenous (IV) cannula was inserted into the participant’s arm for blood sampling. For MMT involving study drug, participants took the study drug on arrival at the research centre. After 60 minutes, a standard liquid meal (Fortisip Compact, Nutricia, NL) was given. Blood samples for insulin, C-peptide and glucose were taken at 7 defined time points: 0, 15, 30, 35, 60, 90 and 120 minutes. A single sample for Total GLP-1, GIP and glucagon concentrations was taken at time 0. Plasma concentrations of gliclazide were sampled at time 0, 60, and 120 minutes in all participants (n=30); 9 participants were consented to complete a prolonged MMT for further sampling at 4, 8 and 24 hours for gliclazide pharmacokinetic profiling. The end of study was determined as last patient last visit.

## Materials

### Study Drugs

Gliclazide 40mg tablets were sourced from Alliance Pharmaceuticals (UK) and halved by Tayside Clinical Trials Pharmacy. The DPP4i was sitagliptin in the form of Januvia 100mg tablets (Merck Sharp and Dohme Ltd, UK). A 100mg once daily dosing schedule was chosen as pharmacokinetic studies in healthy individuals showed a more consistent DPP4 inhibition profile over a 24-hour period. In comparison, a 50mg dose only provided 12-hours of consistent DPP4 inhibition (12).

### Liquid Meal

The liquid meal comprised 160ml of Fortisip Compact, the nutritional content for the given volume was: 184 kilocalories, protein 15.36 grams, carbohydrate 47.52 grams, fat 14.88 grams, nil fibre.

### Continuous Glucose Monitors

The Freestyle Libre Pro Flash Continuous Glucose Monitoring System was used throughout study (Abbott).

## Blood Collection

All blood collection was performed utilising BD Vacutainer systems (Becton, Dickinson and Company, NJ, USA). Samples were iced following collection and centrifuged immediately in accordance with recommended guidance from receiving laboratories.

## Laboratory Analyses

### Insulin and C-Peptide

Analysis of insulin and C-peptide was performed by Clinical Chemistry, Royal Devon, and Exeter Hospital – 602 modules Cobas 8000 automated platform using sandwich chemiluminescence immunoassay (Elecsys insulin, Belgium)

### Glucose

Glucose analysis was performed by NHS Tayside Blood Sciences at Ninewells Hospital utilising Siemens ADVIA Chemistry, Glucose Hexokinase_3 Concentrated Reagents (UK).

### Glucagon

Glucagon analysis was performed by the Immunoassay Core Biomarker Laboratory, University of Dundee, utilising EMD Millipore glucagon radioimmunoassay kit (Merck, Billerica, MA, USA).

### Incretins

Total GLP-1 and GIP analyses were performed by the Immunoassay Core Biomarker Laboratory, University of Dundee, utilising MSD metabolic assay Total GLP-1 and GIP assay (MSD, MD, USA).

### Gliclazide

Gliclazide analysis was performed by the Biomarker and Drug Analysis Core Facility, University of Dundee utilising a uniquely developed gliclazide quantification method in human plasma by liquid chromatography separation, and tandem mass spectrometry analysis.

## Data and Statistical Analyses

### Study Outcomes

The statistical analysis is included in Supplementary Information. The primary outcome was the change in glucose sensitivity at MMT. Secondary outcomes included: the effect of treatment on parameters of beta-cell function, biochemical parameters (glucose, insulin, c-peptide, and incretin hormones) and the pharmacokinetic profile of low dose gliclazide. The frequency of blood sugar levels <3mmol/l on CGM, and the effect of KCNJ11 (E23K) genotype and gender on change in glucose sensitivity with drug treatment were also evaluated.

### Randomisation

Participants were randomised to intervention order using an unblinded web-based randomisation software (www.randomisation.com). A copy of the randomisation plan was stored in the research centre, Tayside Clinical Trials Pharmacy, and the site file.

### Power

Based on previous data in T2DM, the standard deviation of the difference in AUC glucose between placebo and vildagliptin treatment was 125mmol/l over 240 minutes (13). With 30 patients, the study would have 80% power (*p*=0.05) to detect a difference of 1/3 of that seen with vildagliptin alone compared with placebo. Similarly, the power would be enough to detect approximately 50% of the difference in AUC_INSULIN_:AUC_GLUCOSE_ ratio seen comparing vildagliptin and placebo.

### Modelling

Two models were developed, a linear mixed effects model and a generalised additive model, both of which considered the hierarchical nature of the study design of three levels: treatment (n=4), participant (n=30) and time within the MMT (n=7 time points per MMT). Participants were randomised to block order and the MMT took place on either day 14, 28, 42 or 56 from the start of study.

#### Linear Mixed Model with Random Effect

For the primary outcome a linear mixed effects model was applied (14) with glucose sensitivity as the dependent variable. Synergy was evaluated via post-hoc pairwise comparisons between treatments. In this model, treatment intervention was considered a fixed effect, as was time within the MMT. Inter-subject variability, block randomisation and day of MMT were considered as random effects. Finally, ⍰ accounted for all other random effects. All assumptions of linear mixed effect model residuals were checked for deviation from homoscedasticity or normality.

#### Generalised Additive Model

As the time course of the insulin, c-peptide, incretin, and glucagon response across the MMT were considered as non-linear parameters, a generalised additive model was developed (15). In this analysis, “*Treatment*” was considered as a fixed effect whereas “*Time*” was considered as a fixed, non-linear effect. The model applied smoothing parameters to random effects of inter-subject variation, block randomisation and day. The model fit was checked using the “*mgcv*” package in R (15).

### Parameters of Beta Cell Modelling

Beta cell function was assessed using a previously described model (16, 17)(18), designed to analyse the MMT tests. The model describes the relationship between insulin secretion and glucose concentration by means of a dose-response function relating the two variables and an early secretion component. The dose-response is characterised by its average slope, termed glucose sensitivity, and early secretion by a parameter denoted as rate sensitivity, a marker of early phase insulin release. The dose response function is modulated by a time-varying potentiation factor, which accounts for effects of sustained hyperglycaemia and incretins. The potentiation factor excursion was calculated as the ration between the values at the end of the 2-h OGTT and at baseline.

### Data Presentation

Model results are presented in tables of estimates (mean (95% confidence intervals)), standard error, test statistic and *P*-value. A significant *P*-value result provided in the “*Control*” arm represents that the intercept is significantly different to 0. *P*-values for treatment interventions demonstrate statistical significance versus control.

### Statistical Software

Data were managed utilising Microsoft Excel as part of Microsoft Office 365 Pro Plus Version 1908 (Build 11929.20708). Statistical analysis and graphical presentation were performed in R (19).

## Results

### Baseline Characteristics

Study recruitment ran from September 2019 to September 2020. All study activity was paused due to COVID-19 pandemic between March and August 2020. Two participants withdrew consent for study immediately after screening in March 2020 due to the COVID-19 pandemic; these participants were replaced. One participant withdrew prior to last MMT due to circumstances unrelated to study; data until point of withdrawal were retained as per study consent.

Baseline characteristics of the study cohort (n=30) were representative of SU users in the Tayside region (Table 1). In a sub-analysis by gender, male participants had lower BMI (median (LQ, UQ)) (Male 30.5 (25, 33) vs Female 39 (31, 41) kg/m (*p*<0.001)) and were older (Male 67.5 (64, 71) vs Female 59 (54, 66) years, (*p* = 0.02). The most common concomitant medications were metformin (n=27), statins (n=26), proton pump inhibitors (n=13) and selective serotonin reuptake inhibitors (11).

**Table 1:** Baseline Characteristics (Median (Lower Quartile, Upper Quartile)). 30 Participants were analysed for the primary outcome of study. 9 Participants completed prolonged mixed meal tolerance tests for 24-hour sampling for low dose gliclazide pharmacokinetic profiling.

| Phase of Study | Number of Participants | Gender (M/F) | Pre-Existing Treatment (Diet/Metformin) | Age (Years) | Body Mass Index (Kg/M<sup>2</sup>) | Body Surface Area (M<sup>2</sup>) | HbA1c (mmol/mol) | Duration of Diabetes (Years) |
| --- | --- | --- | --- | --- | --- | --- | --- | --- |
| Full Study | 30 | 16/14 | 3/27 | 66<br>(57, 70) | 32.4<br>(28.40) | 2.1<br>(1.9, 2.3) | 54<br>(48, 62) | 6.5<br>(4.8, 10.0) |
| Pharmacokinetic Phase | 9 | 3/6 | 3/6 | 66 (64, 72) | 30.7<br>(27, 38) | 2.1<br>(1.7, 2.4) | 50<br>(48, 60) | 5<br>(5, 10.5) |

Adverse events included one episodes of symptomatic hypoglycaemia (BM 3.3 mmol/l) which occurred on DPP4-inhibitor monotherapy. There were 14 occurrences of detachment of CGM sensors which were documented as adverse events. Sensors were replaced on the next working day following report to the study team.

### Primary Outcome: change in glucose sensitivity with treatment

Linear mixed model parameters of beta cell function are summarised in Table 2. The plot of glucose sensitivity suggests additive effect of treatment (Figure 2A). However, the linear mixed model estimates only show difference in glucose sensitivity with SUDPP4i compared to baseline (*p*=0.04) (Table 2).

**Table 2:**
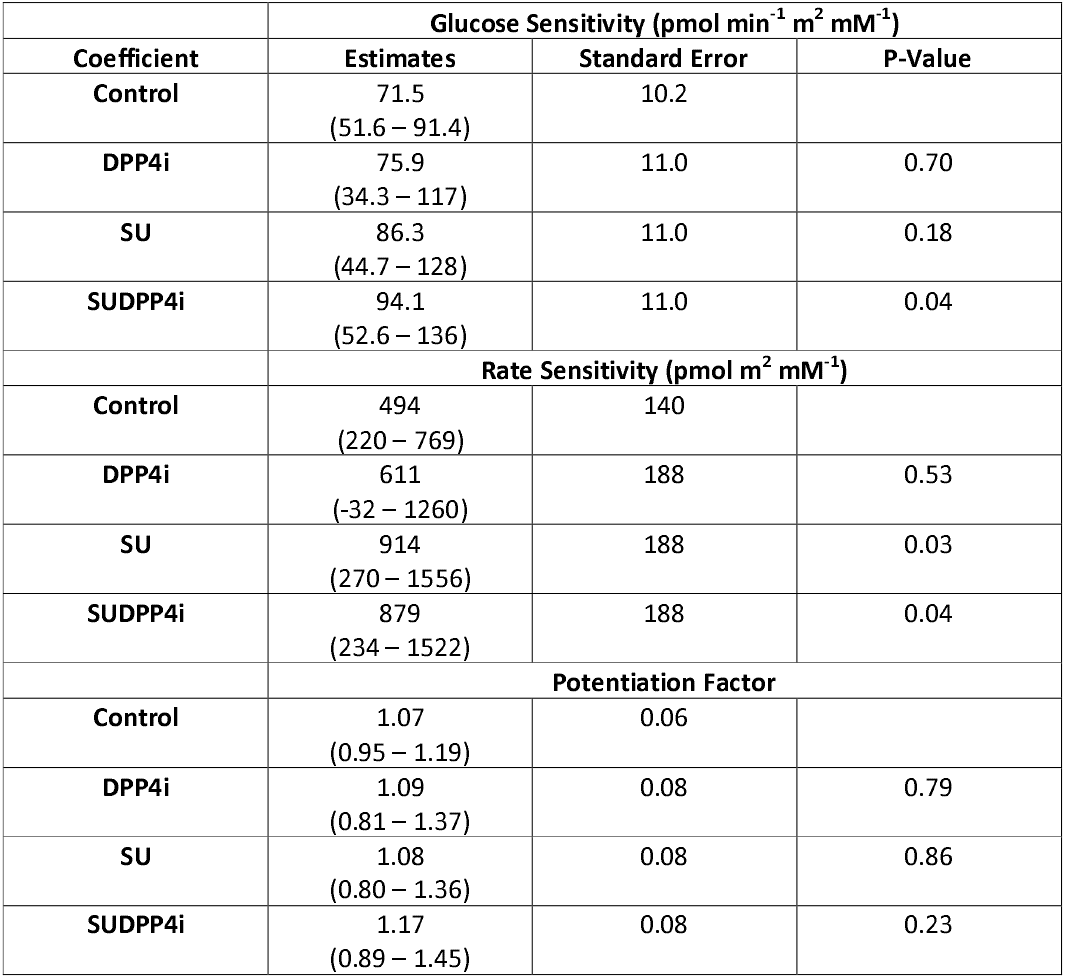
Linear mixed effects modelling results of parameters of beta cell function. Estimates are shown as mean (95% confidence intervals). All values are rounded to 3 significant figures. *P*-values for treatment interventions demonstrate statistical significance versus control.

**Figure 2.**
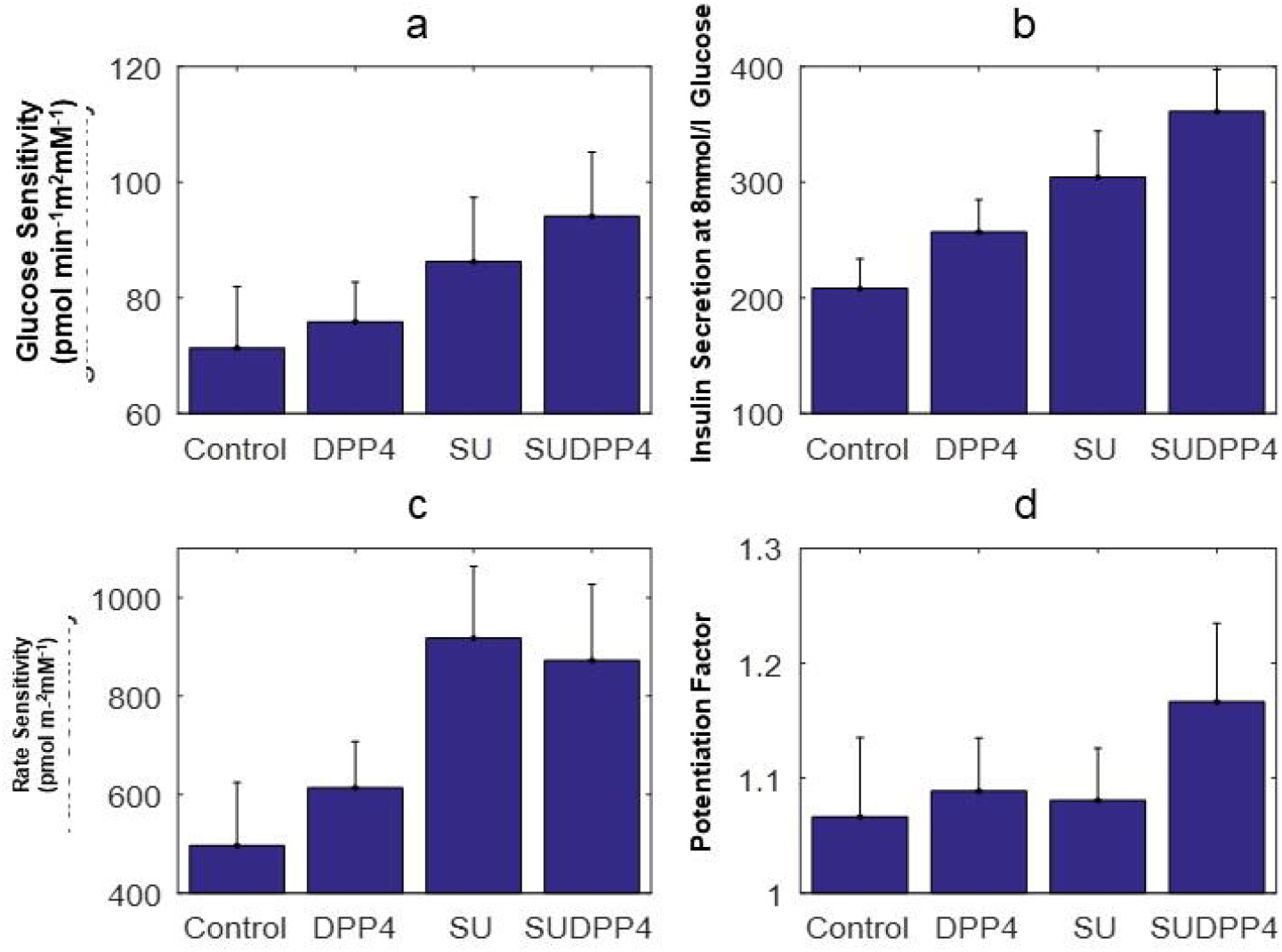
a) Glucose sensitivity b) Insulin Secretion at 8mmol/l glucose c) Rate Sensitivity d) Potentiation Factor by treatment (Mean (SEM))

Figure 3 shows the model-determined relationship between insulin secretion and glucose concentration in each of the four intervention groups. A progressive increase in slope is observed across the treatments, representative of the corresponding increase in glucose sensitivity. A left shift is noted in favour of combination treatment in both glucose sensitivity and insulin secretion at 8mmol/l (Figure 2B), demonstrating augmentation of beta cell function at lower glucose concentrations.

**Figure 3.**
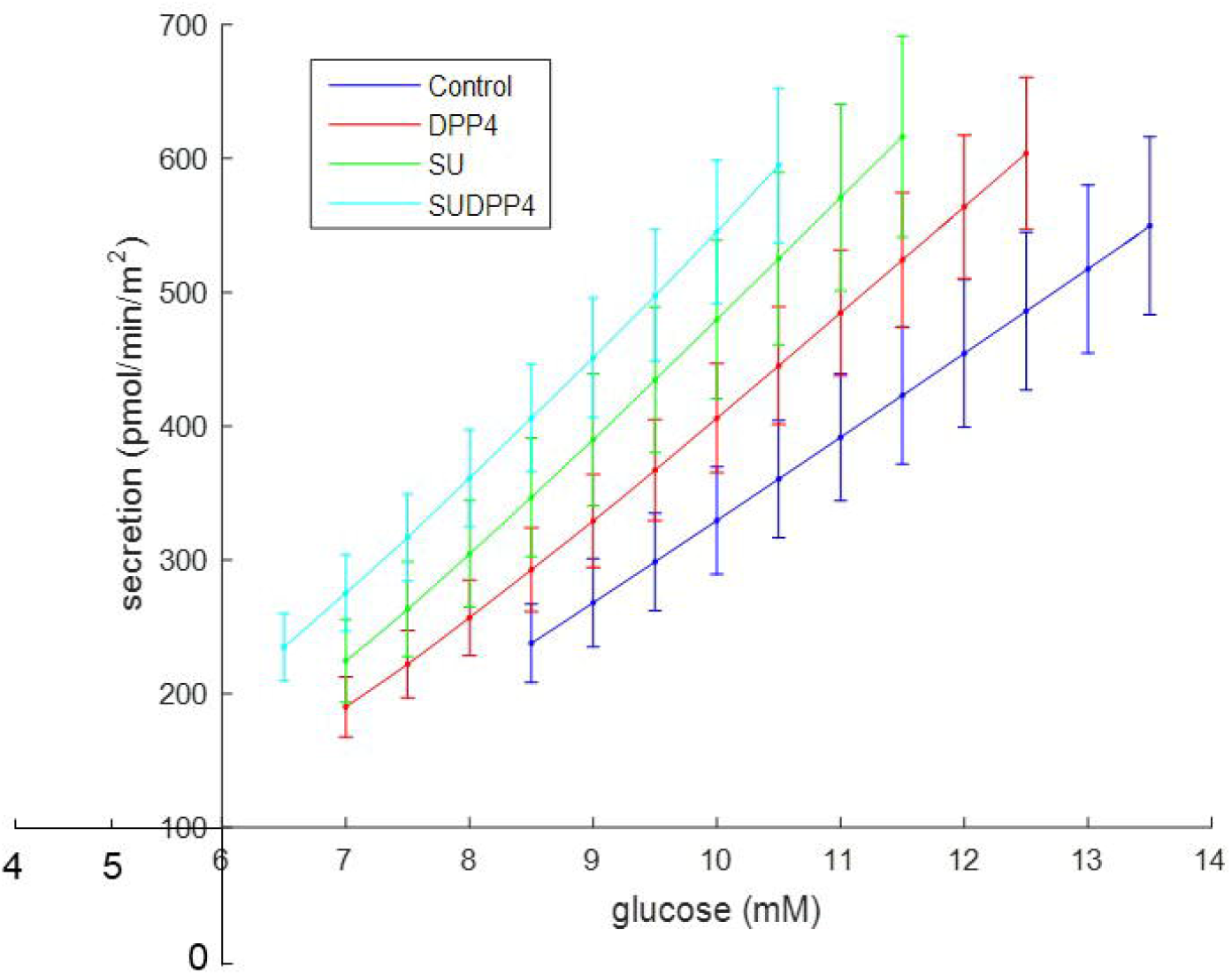
Dose response by treatment (Mean (SEM))

### Secondary Outcomes

Rate sensitivity, which is a marker of early insulin release, was augmented by both gliclazide interventions (Figure 2C, Table 2). This is expected, as gliclazide is a secretagogue influencing early insulin secretion. Predictably, this effect was not further augmented by the combination with DPP4i.

There was no difference in potentiation factor ratio (Table 2). The trend towards an increase in SUDDP4i suggests that had the MMT been prolonged (Figure 2D), difference may have been observed in late phase potentiation as in our previous study on the incretin effect, which lasted four hours (11).

### Glucose

The mean fasting and mean glucose AUC were reduced in all treatment groups compared with control (Figure 4). Linear mixed modelling outcomes showed additive effect in terms of difference between estimates of both DPP4i and SU groups versus SUDPP4i (Table 3). SUDPP4i reduced mean glucose AUC compared with both treatments as monotherapy.

**Figure 4.**
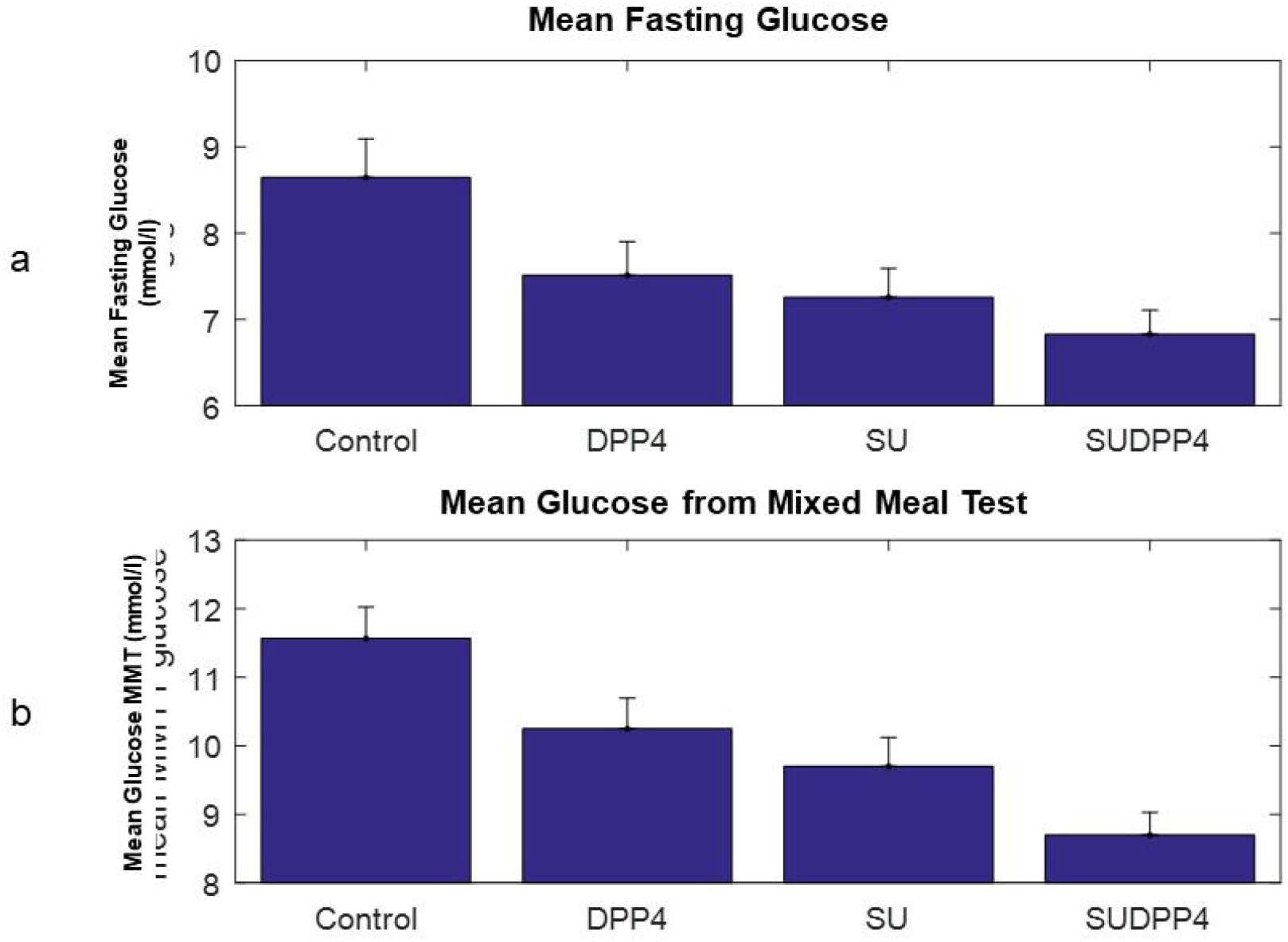
Mean (SEM) a) Fasting Glucose b) Glucose from Mixed Meal Test

**Table 3:** Summary of linear mixed model outcomes for glucose and generalised additive model outcomes for insulin, and C-peptide from mixed meal tolerance test. Estimates are Mean (95% Confidence interval). All values are rounded to 3 significant figures. *P*-values for treatment interventions demonstrate statistical significance versus control.

| Coefficient | Mean Fasting Glucose (mmol/l) |  |  | Mean Glucose from AUC (mmol/l) |  |  |
| --- | --- | --- | --- | --- | --- | --- |
|  | Estimates (95% CI) | Std Error | P-Value | Estimates (95% CI) | Std Error | P Value |
| Control | 8.59<br>(7.88 – 9.31) | 0.37 |  | 11.5<br>(10.7 – 12.3) | 0.42 |  |
| DPP4i | 7.51<br>(7.06 – 8.68) | 0.23 | <0.001 | 10.25<br>(8.84 – 11.7) | 0.30 | <0.001 |
| SU | 7.25<br>(6.09 – 8.42) | 0.23 | <0.001 | 9.7<br>(8.29 – 11.1) | 0.30 | <0.001 |
| SUDPP4i | 6.83<br>(5.6 – 8) | 0.23 | <0.001 | 8.7<br>(7.29 – 10.1) | 0.30 | <0.001 |
| Coefficient | Fasting Insulin (pmol/l) |  |  | Fasting C-Peptide (pmol/l) |  |  |
|  | Estimates (95% CI) | Std Error | P-Value | Estimates (95% CI) | Std Error | P Value |
| Control | 170<br>(117 – 224) | 27.1 |  | 1503<br>(1271 – 1735) | 118 |  |
| DPP4i | 137.17<br>(55.2 – 219) | 14.8 | 0.03 | 1370<br>(1016 – 1725) | 62.7 | 0.04 |
| SU | 165.46<br>(83.5 – 199) | 14.8 | 0.76 | 1495<br>(1141 – 1620) | 62.7 | 0.9 |
| SUDPP4i | 182.24<br>(99.6 – 265) | 15.1 | 0.42 | 1615<br>(1261 – 1969) | 62.7 | 0.07 |
| Coefficient | Incremental AUC Insulin (nmol/l/min<sup>-1</sup>) |  |  | Incremental AUC C-Peptide (nmol/l/min<sup>-1</sup>) |  |  |
|  | Estimates (95% CI) | Std Error | P-Value | Estimates (95% CI) | Std Error | P Value |
| Control | 41.1<br>(30.4 – 51.7) | 5.39 |  | 144<br>(111 – 177) | 16.8 |  |
| DPP4i | 43.0<br>(18.2 – 34) | 7.56 | 0.71 | 159<br>(120 – 131) | 19.8 | 0.45 |
| SU | 42.0<br>(16.4 – 67.6) | 7.55 | 0.90 | 151<br>(112 – 224) | 19.8 | 0.72 |
| SUDPP4i | 44.3<br>(18.7 – 70.1) | 7.58 | 0.66 | 154<br>(81.2 – 226) | 19.8 | 0.65 |

### Insulin and C-Peptide

The generalised additive model estimates showed no effect of treatment on either incremental AUC (iAUC) insulin or C-peptide (Table 2). However, when interpreted in context of significant glucose reduction, this would suggest an overall improvement in beta cell function (Table 3).

### Incretins

As our previous work found that low dose gliclazide had no impact on dynamic endogenous incretin or glucagon secretion (11), only fasting measurements were performed. Generalised additive model estimates showed that SU reduced fasting GLP-1, GIP, and glucagon concentrations (Supplementary Information Table 1).

### Gliclazide Pharmacokinetics

Study drugs were administered 60 minutes prior to the start of the MMT. Mean gliclazide concentrations (mean (SD)) (ng/ml) were SU 662 (408), SUDPP4i 603 (355) respectively (*p* = 0.31) and maximum concentrations were 749 (433) and 645 (397) ng/ml (*p*=0.01) in the SU and SUDPP4i meal tests, respectively. Combination treatment did not affect the 24-hour profile of gliclazide: mean gliclazide AUC 660 (328) and 535 (223) ng/ml (*p*=0.1), maximum concentration 770 (328) and 793 (413) (*p*=0.8). Trough concentrations of gliclazide were 370 (183) and 343 (183) ng/ml in the SU and combination groups, respectively.

### Continuous Glucose Monitoring

Blood glucose <3mmol/l was considered as biochemically significant hypoglycaemia in line with the European and American joint position statement (21). Linear mixed effects modelling of time in range (TIR) <3mmol/l on CGM (%) was unaffected: Control 1 (2-4), DPP4i 2 (3-6), SU 1 (0-4) (*p*=0.64). Only treatments involving SU increased TIR 3-10mmol/l (%) versus control: Control 67.4 (56.6 – 78.2), DPP4i 64.5 (45.6 – 83.74), SU 71.83 (52.59 – 17.25), SUDPP4i 68.4 (66.16 – 85.83) (*p*<0.001 SU & SUDPP4i versus control).

### Effect of *KCNJ11* (E23K) Genotype

The study cohort included 12 EE, 5 KK and 12 EK heterozygotes, which is slightly higher than the 34 – 48% minor allele frequency reported for Caucasian populations (22). Linear mixed model estimates showed additive effect of treatment on glucose parameters in EE homozygotes only (Supplementary Information Table 2) and fasting insulin and C-peptide in those carrying the K-allele. Plots of the glucose sensitivity suggest a steeper slope in K-allele carriers (Figure 5); however, the linear mixed model estimates were not statistically significant.

**Figure 5.**
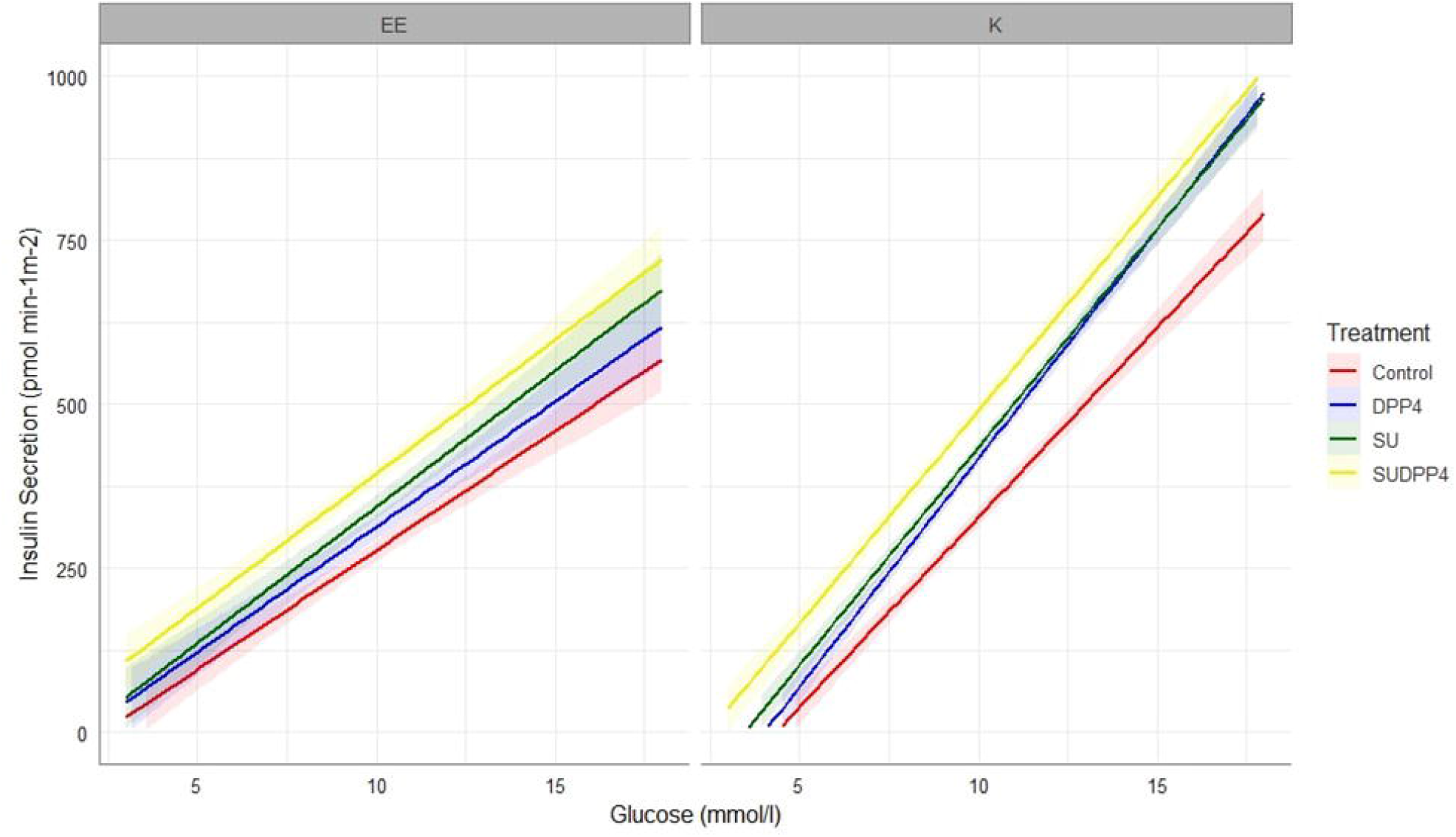
Dose response to treatment by *KCNJ11 E23K* genotype (Mean (SEM))

### Effect of Gender

As previous literature had suggested differing response to SU by sex and BMI (23), the model was adjusted for this interaction. Sub analysis of glucose sensitivity and the dose response revealed a potent additive effect in male participants, which was not observed in female participants (Figure 6). Linear mixed modelling estimates suggest that women respond better to DPP4is, which has not been previously documented, with no difference in glucose lowering effect between DPP4i and SU. In contrast, men showed a greater response to interventions involving SU, including between SU and combination treatment (Supplementary Information Table 3).

**Figure 6.**
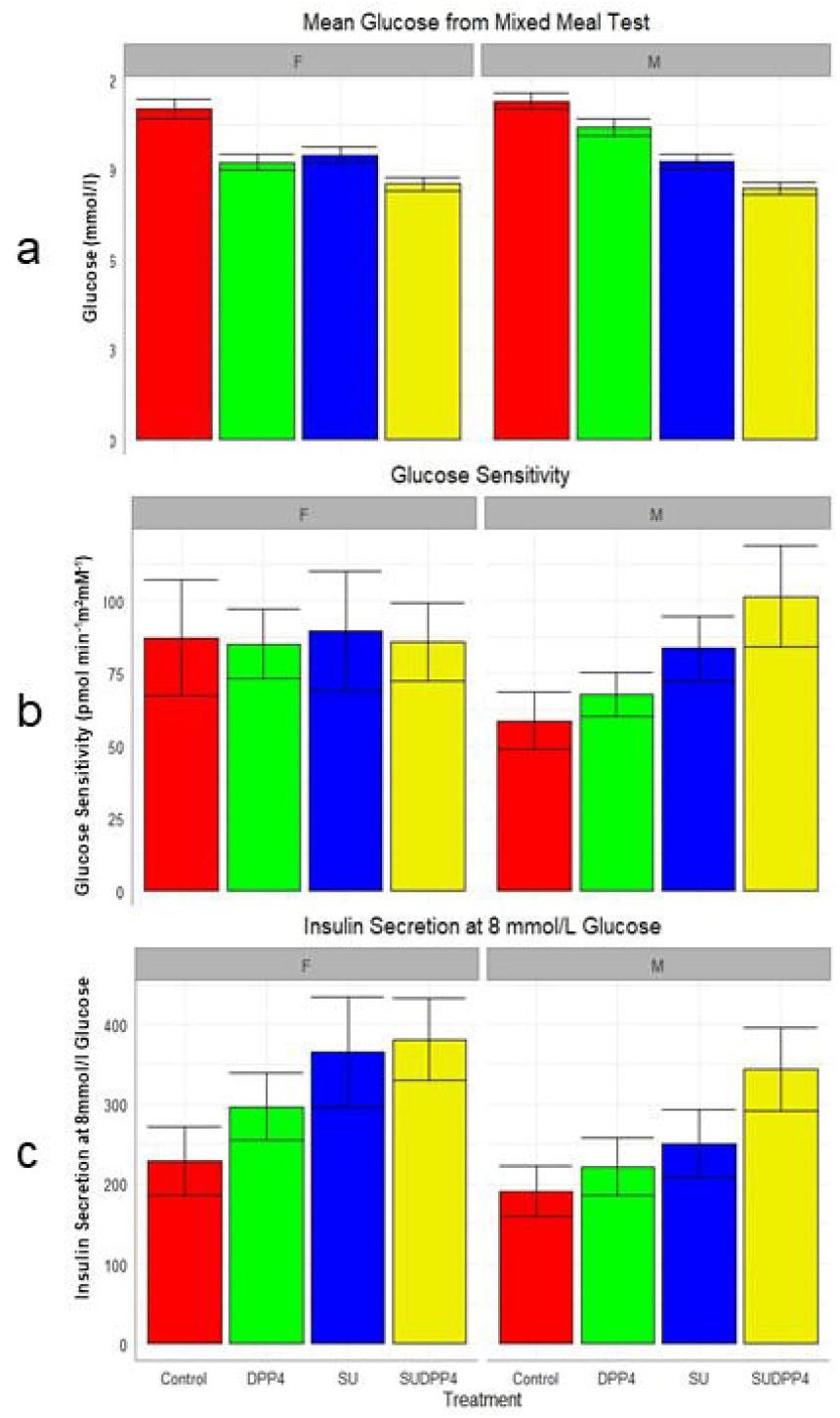
Gender Differences in Treatment Response (Mean (SEM)) a) Glucose from Mixed Meal Test b) Glucose Sensitivity c) Insulin Secretion at 8mmol/L Glucose

## Discussion

The study describes the effect of low dose SU as monotherapy, or in combination with a DPP4i on parameters of beta-cell function in response to a standardised meal with four different interventions: control, 100mg sitagliptin, 20mg gliclazide or both. Our results show that combination treatment with SU and DPP4i enhanced glucose control and beta-cell function, without hypoglycaemia on CGM which supports that this combination could have potential use as an effective, low-cost treatment. However, the observed increase in glucose sensitivity with combination treatment was not greater than the sum of the monotherapy responses.

### Low dose sulphonylureas are potent glucose lowering agents whilst avoiding hypoglycaemia

In this study, we show that 20mg of gliclazide has potent glucose lowering potential which is further enhanced by the addition of a DPP4i whilst avoiding hypoglycaemia. A 2.8 (0.305) mmol/l (mean (SEM)) reduction in mean glucose from AUC was observed with combination treatment versus control. Strikingly, 20mg of gliclazide as monotherapy was as potent as 100mg sitagliptin (mean glucose reduction 1.8 mmol/l (0.3), 1.3 (0.3) mmol/l *p*=0.27 gliclazide vs sitagliptin respectively). A meta-analysis of studies combining standard dose SU with DPP4i have shown a 50% increased risk in hypoglycaemia in the first 6 months of treatment (24). We used 20mg gliclazide in this study, which showed no difference in the frequency of hypoglycaemic events between treatments.

We propose mechanistically that low dose SU do not cause hypoglycaemia due to their effect on the K_ATP_ channel open state, similar to their mechanism observed in *KCNJ11* neonatal diabetes mellitus (NDM) (25). In *KCNJ11* NDM, high dose glibenclamide is required to promote insulin secretion and successful transition off insulin (7). Even at these high doses the mutant K channels do not shut completely, resulting in a beta-cell resting membrane potential that is sub-threshold for insulin release but primed for other stimuli such as incretins. In T2DM, where there are only minor defects in K_ATP_ channel function, normal doses of SU fully close the K_ATP_ channels resulting in insulin secretion despite normal or low glucose. Our findings suggest that a very low dose of SU in T2DM achieves a similar partial closure of the K_ATP_ channels as seen for high dose SU in NDM, working primarily to prime the beta-cell to other secretagogues such as the incretins or amino acids, resulting in glucose regulated insulin secretion and no insulin secretion in the presence of normal or low blood glucose. Therefore, it could be possible to achieve the glycaemic benefits of SU, whilst minimising negative attributes.

### Combination low dose sulphonylurea and DPP4i heighten parameters of beta cell function with additional effect on glucose lowering

Modelling of beta cell function showed progressive augmentation of the slope of the dose-response (glucose sensitivity) in favour of SUDPP4i, however, although there was clear additive effect, there was no evident synergy as hypothesised. The relationship of the dose-response is presented in Figure 3; there are two parameters characterising this relationship. The first is glucose sensitivity, and the second is insulin secretion at fixed glucose concentration, which is equivalent to an intercept. It may be that the left shift in the dose-response may at least in part be independent of glucose sensitivity, however in this study significant additive effect is also seen in this parameter. It can be postulated that SU in this instance are enhancing insulin section, but maintaining glucose dependence, thus avoiding hypoglycaemia, as supported by our CGM findings. The impact of gliclazide on glucose sensitivity has been previously suggested in rat models (26, 27) and in healthy human participants (28), albeit at high dose.

### Gliclazide pharmacokinetics

This study explored the 24-hour plasma concentration profile of 20mg standard release gliclazide, observing trough concentrations of ∼370ng/ml, mean plasma concentrations of 500 - 600ng/ml and peak of 908 ng/ml. For comparison, an 80mg dose of gliclazide generates peak plasma concentrations of between 3000 – 5000ng/ml (29). Interestingly the plasma concentrations observed in this study are only a little lower than those documented for 30mg gliclazide MR: trough 472, mean 800 and maximum concentrations of 1100 ng/ml, respectively. A multi-centre double-blind RCT compared the efficacy and safety of gliclazide MR vs glimepiride, in a cohort which included those at higher risk of SH (>65 years and renal impairment). Both groups achieved HbA1c reduction of 1.0% with fewer hypoglycaemic events with gliclazide MR than glimepiride (3.7 vs 8.9%) (30), which suggests that low dose modified release preparations may be preferential in terms safety with similar cost and efficacy to standard release SU.

### Response by Genotype

In this study K allele (diabetes-risk allele) carriers showed higher fasting glucose, reduced insulin secretion and beta cell function. Differences were observed in terms of plasma glucose and beta-cell function in carriers of the K allele, but there was no difference in the response to treatment. Plots of the dose response suggest that K-allele carriers have lower fasting insulin secretion, but in the control group there was a greater slope of glucose sensitivity than EE homozygotes, similar to previous literature (31). However, the difference in slopes with gliclazide treatment was more pronounced in EE homozygotes suggesting that the 20mg dose may not be high enough to sufficiently close the K_ATP_ channels in K allele carriers to allow the amplifying pathway to operate. This is supported by a previous study which suggested that K allele carriers require higher doses of SU to achieve glucose reduction, and even higher doses in KK homozygotes (32). A dose-response by genotype study would be required to fully investigate this effect.

### Gender differences in response

It is interesting that the additive pattern of glycaemic reduction and increase of beta-cell function with SU or in combination with a DPP4i is only seen in men, but not women. However, sub analysis shows the additive effect on insulin secretion at 8mmol/l glucose is preserved in women, which does suggest effect independent of glucose sensitivity in this instance. These differences in physiological responses by gender mirror findings in large cohort studies (23). In an analysis of subgroups of BMI and gender in patients (n=22,379) starting SU or TZD in the UK Clinical Practice Data Research Datalink (CPRD), non-obese males (BMI <30) had a 3.3 mmol/mol better response to SU than TZD (p<0.001), these findings were replicated in the ADOPT study (first-line treatment) (33) and observed in the RECORD study (34). These studies support that there is a sex difference in SU response, and our study provides some physiological insights into these differences. Possible explanations include that the women had higher BMI, although adjusting for BMI does not remove the sex difference (data not shown); or a sex difference in incretin physiology. The ADDITION-PRO study reported that women have greater increased serum GLP-1 concentrations following OGTT than men, even after adjustment for BMI (35). Further studies are warranted to further investigate this.

## Limitations

The main limitation in this study is the wide variability in beta-cell response within a small cohort, which limits power and ability to perform further sub-analysis. However, the population would reflect those who would merit second-line intensification in real-world medicine. This open-label physiological study design was adopted as further proof-of-concept prior to undertaking a formal randomised controlled trial (RCT) The ideal study design would have been a double-blinded RCT, but this was not feasible at this stage.

The advantage of beta-cell modelling is the ability to model static and dynamic parameters of beta-cell function, beyond traditional measures of glucose and insulin secretion. However, as a more complicated procedure, involving multiple parameters it may add some estimation error.

## Conclusion

We have shown that low dose sulphonylureas are potent glucose lowering agents, which increase the beta-cell dose response to glucose without increasing hypoglycaemia. This response is further augmented in the presence of a DPP4 inhibitor, although we did not see synergy with this combination. A formal randomised controlled trial of the efficacy and safety of a low dose sulphonylurea in combination with a DPP4 inhibitor is warranted as a combination treatment may allow modernisation of two cheap, effective treatments of T2DM with considerable potential for pharmacoeconomic benefit worldwide.

## Acknowledgements

The authors would like to thank the study team of The Clinical Research Centre, Ninewells Hospital and Medical School, Dundee: H Loftus, L Cabrelli, G Wilkie, G Kiddie, D Pankhurst and C Shearer. Thanks to T McDonald and R Nice of the Department of Clinical Chemistry, University of Exeter for insulin and c-peptide measurements. J Burns of Ninewells Hospital and Medical School, Dundee for measurement of glucose concentrations, J Huang and the Department of Biomarker and Drug Analysis, University of Dundee for the development of the gliclazide mass spectrometry method and the Immunoassay Core Biomarker Facility, University of Dundee for incretin and glucagon measurement.

## Data Availability

Some or all datasets generated and/or analysed during the current study are not publicly available but are available from the corresponding author on reasonable request.

## Contribution Statement

RLMC and ERP conceived, designed and acquired study data. RLMC, KB and ERP designed the statistical analysis plan. RLMC undertook statistical analysis with advice from KB, ERP and AM. AM and AT conducted beta-cell modelling analysis and interpretation. RLMC and EP wrote the initial drafts of the manuscript, these drafts were revised for important scientific content by all authors. All authors gave final approval of the version to be published. ERP is the guarantor of this work.

