## Supplementary Information for "Combination low dose sulphonylurea and DPP4 inhibitor have potent glucose lowering effect through augmentation of beta cell function without increase in hypoglycaemia: a randomised crossover study"

### 1 Supplementary Table 1: Summary of generalised additive model outcomes for GLP-1, GIP, and glucagon from mixed meal tolerance test.

### 2 Supplementary Table 2: Summary of linear mixed effects model outcomes for glucose, and generalised additive model outcomes for insulin, and C-peptide from mixed meal tolerance test by *KCNJ11* genotype

### 3 Supplementary Table 3: Summary of linear mixed effects and generalised additive model outcomes from mixed meal tolerance test by gender

### 4 Statistical Analysis Plan

|  | **GLP-1 (pg/ml)** | | |
| --- | --- | --- | --- |
| **Coefficient** | **Estimates** | **Standard Error** | **P-Value** |
| **Control** | 24.1  (18.3 – 29.9) | 3.0 |  |
| **DPP4i** | 21.1  (16.5 –25.7) | 2.3 | 0.2 |
| **SU** | 17.5  (12.8 – 27.8) | 2.3 | 0.004 |
| **SUDPP4i** | 21.8  (11.38 – 21.7) | 2.4 | 0.337 |
|  | **GIP (pg/ml)** | | |
| **Control** | 176  (132 – 219) | 21.9 |  |
| **DPP4i** | 105  (53.4 – 157) | 26.4 | 0.01 |
| **SU** | 105  (53.7 – 157) | 26.6 | 0.01 |
| **SUDPP4i** | 143  (91 – 156) | 26.7 | 0.22 |
|  | **Glucagon (pmol/l)** | | |
| **Control** | 9.95  (7.3 – 12.6) | 1.33 |  |
| **DPP4i** | 10.3  (8.63 – 11.9) | 0.82 | 0.72 |
| **SU** | 7.82  (6.21 – 9.43) | 0.82 | 0.01 |
| **SUDPP4i** | 11.5  (7.26 – 15.8) | 0.83 | 0.23 |

**Supplementary Table 1**: Summary of generalised additive model outcomes for GLP-1, GIP, and glucagon from mixed meal tolerance test. Estimates are Mean (95% Confidence interval). *P*-values for treatment interventions demonstrate statistical significance versus control.

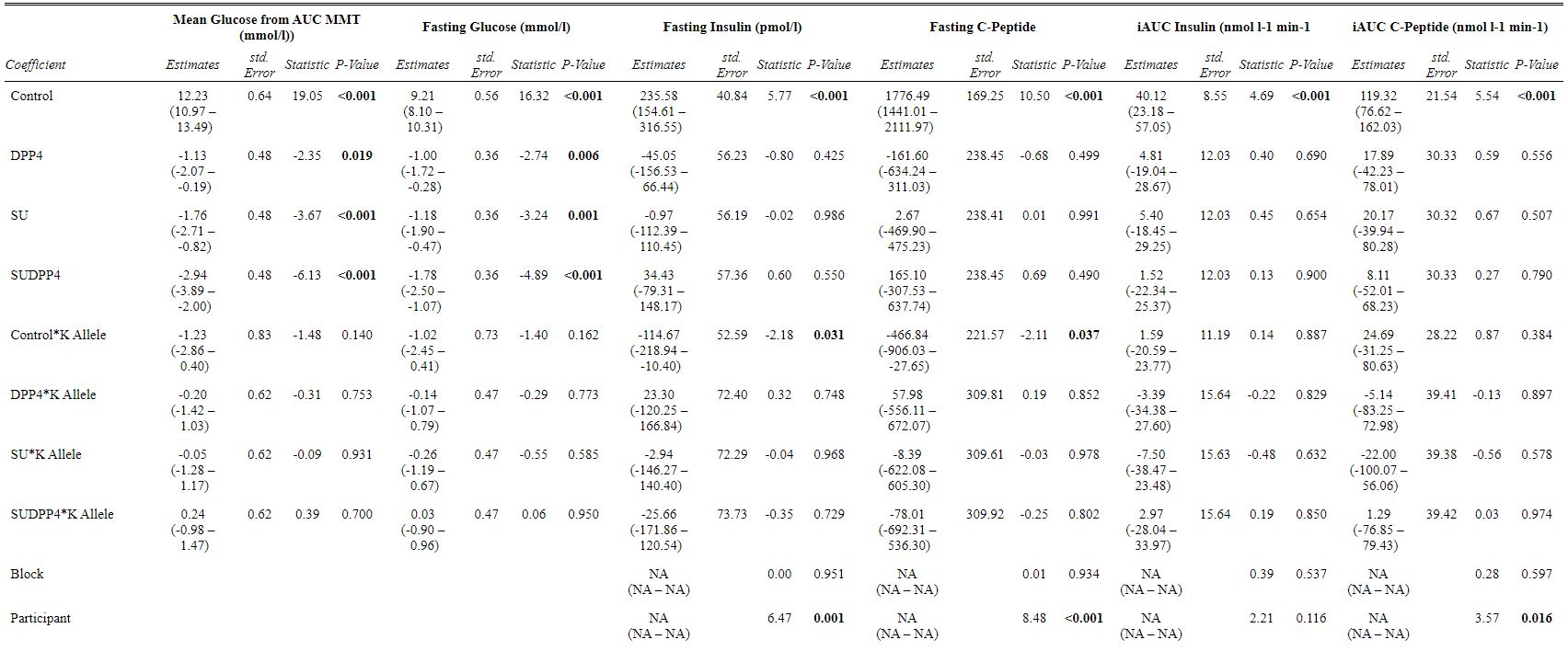

**Supplementary Table 2:** Summary of linear mixed effect model outcomes for glucose, and generalised additive model outcomes insulin, and C-peptide from mixed meal tolerance test by *KCNJ11* genotype (EE Blue, KK Red). Estimates are Mean (95% Confidence Interval). *P*-values for treatment interventions demonstrate statistical significance versus control.

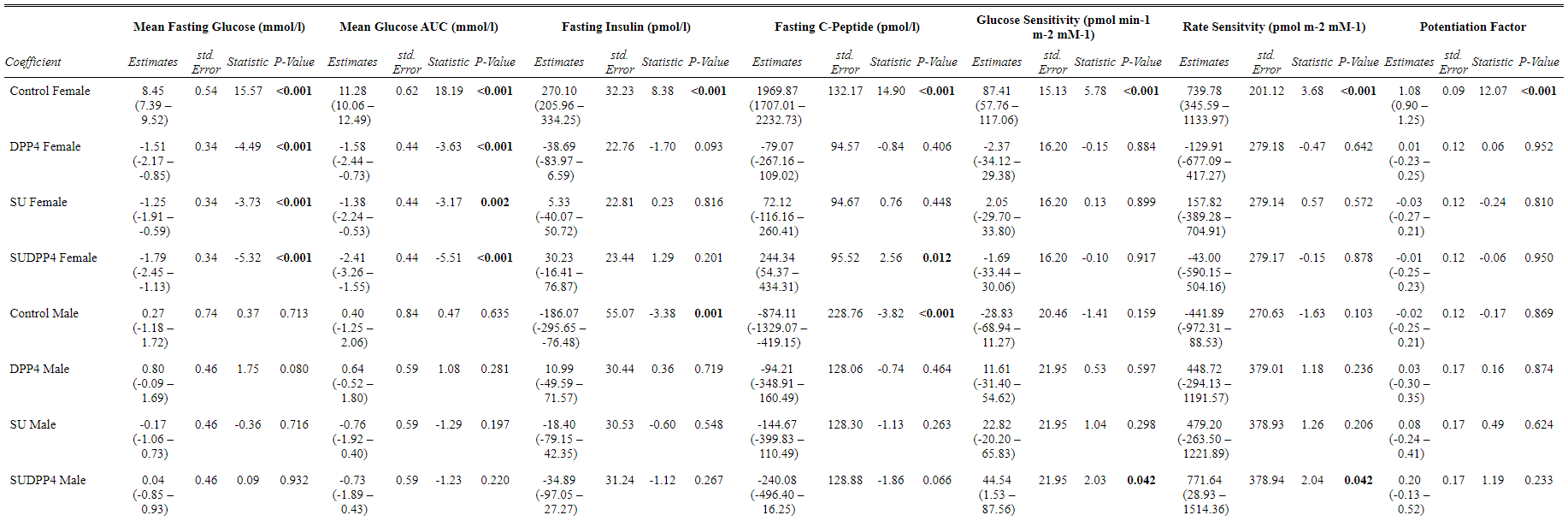

**Supplementary Table 3**: Summary of linear mixed effect and generalised additive model outcomes from mixed meal tolerance test by gender (Male Blue, Female Red). Estimates are Mean (95% Confidence Interval).

SSS Study

Statistical Analysis Plan

| STUDY FULL TITLE | Study of Sulphonylurea Synergy with DPP4 Inhibitors |
| --- | --- |
| Clinical Trials.gov Number | NCT04192292 |
| R&D Number | 2018DM13 |
| REC Number | 19/ES/0092 |
| IRAS ID | 266525 |
| SAP VERSION | 4.0 |
| SAP VERSION DATE | 16/11/2020 |
| TRIAL STATISTICIAN | Dr Khaled Bedair |
| TRIAL CHIEF INVESTIGATOR | Professor Ewan Pearson |
| TRIAL PRINCIPAL INVESTIGATOR | Dr Ruth Cordiner |
| SAP AUTHOR | Dr Ruth Cordiner |

### 6 SAP Signatures

I give my approval for the attached SAP entitled SSS dated 17/11/2020

**Chief Investigator**

Name:

Signature:	                                               
Date:

**Statistician**

Name:

Signature:	                                               
Date:

### 7 Contents

### 8 Abbreviations and Definitions

| List of Abbreviations | |
| --- | --- |
| AE | Adverse Event |
| AR | Adverse Reaction |
| CI | Chief Investigator |
| CNORIS | Clinical Negligence and Other Risks Scheme |
| CRF | Case Report Form |
| DM | Diabetes Mellitus |
| DPP-4 | Dipeptidyl Peptidase-4 |
| GCK | Glucokinase |
| GCP | Good Clinical Practice |
| GIP | Gastric Inhibitory Peptide |
| GLP-1 | Glucagon-Like Peptide 1 |
| GoSHARE | Genetics of The Scottish Health Research Register |
| GWAS | Genome Wide Association Study |
| HNF1α | Hepatic Nuclear Factor 1 Alpha |
| HNF4α | Hepatic Nuclear Factor 4 Alpha |
| HOMA | Homeostasis Model Assessment |
| IIGI | Isoglycaemic Intravenous Glucose Infusion |
| IMI-DIRECT | Innovative Medicines Initiative – Diabetes Research on Patient Stratification |
| IMP | Investigational Medicinal Product |
| ISF | Investigator Site File |
| MHRA | Medicines and Healthcare Products Regulatory Agency |
| NICE | The National Institute for Health and Care Excellence |
| NRES | National Research Ethics Service |
| OGIS | Oral Glucose Insulin Sensitivity |
| PI | Principal Investigator |
| PIS | Patient Information Sheet |
| REC | Research Ethics Committee |
| RN | Research Nurse |
| SAR | Serious Adverse Reaction |
| SD | Standard Deviation |
| SDRN | Scottish Diabetes Research Network |
| SIGN | The Scottish Intercollegiate Guidelines Network |
| SOP | Standard Operating Procedure |
| SU | Sulphonylurea |
| SUSAR | Suspected Unexpected Serious Adverse Reaction |
| T2DM | Type 2 Diabetes Mellitus |
| TASC | Tayside Medical Science Centre |
| TMF | Trial Master File |
| UAR | Unexpected Adverse Reaction |

### 9 Introduction

#### 9.1 Brief Summary

The Study of Sulphonylurea Synergy with DPP4 Inhibitors (SSS Study) will establish whether a very low dose of sulphonylurea will have a synergistic role on augmentation of insulin secretion when given in combination with a DPP4 inhibitor. The study will recruit 30 patients with type 2 diabetes controlled with no treatment or metformin monotherapy with an HbA1c <64mmol/mol (<8%).

In this unblinded, randomised crossover study, participants will receive four 14-day intervention blocks: low dose sulphonylurea alone, DPP4 inhibitor alone, low dose sulphonylurea + DPP4 inhibitor or no treatment change.

The primary outcome will be assessed through evaluation of insulin secretion (Incremental Area Under the Curve and Insulin Secretion Rate) at mixed meal test at the end of each treatment block.

Glycaemic variability on continuous glucose monitoring for each intervention block will be evaluated as a secondary outcome. In addition, the primary outcome will be evaluated for KCNJ11 genotype as an additional secondary outcome.

#### 9.2 Purpose of the analyses

- The study will evaluate the difference in parameters of insulin secretion for effect of treatment intervention.
- Sub-analyses will evaluate the primary outcome for baseline screening co-variates as well as KCNJ11 E23K (rs5219) genotype.
- Glycaemic variability glucose during continuous glucose monitoring will be evaluated as a secondary outcome.
- Pharmacokinetics of low dose gliclazide will also be evaluated as a secondary outcome.

#### 1.1 Previous Research Utilised in Analysis Plan Development

##### 1.1.1 Study of Sulphonylurea Synergy with Incretins (LOGIC Study, Clinical Trials.gov NCT03705195)

##### The Study of Sulphonylurea Synergy with Incretins was a proof of concept physiological study involving paired oral glucose tolerance tests and isoglyacemic intravenous glucose infusions in the presence and absence of low dose gliclazide (20mg). It established that low dose sulphonylureas reduces plasma glucose in response to oral glucose load, with concomitant augmentation of the classical incretin effect. Modelling of beta cell function showed that very low plasma concentrations of gliclazide potentiate late phase insulin secretion and increase glucose sensitivity by 50%.

- The incretin effect was defined as the percentage increase in insulin secretion between oral and intravenous glucose. The classical incretin effect was derived as the difference in incremental area under the curve between oral and intravenous interventions utilising the following formula:

Incretin Effect (%) = 100 x (iAUC_OGTT_ – iAUC_IIGI_)

iAUC_OGTT_

- The SSS Study further explores the effect of low dose gliclazide with sitagliptin, a DPP4 inhibitor, which pharmacologically prevents degradation of endogenous incretin hormones through inhibition of the enzyme DPP4.
- The time course of insulin, c-peptide and glucose responses was noted to be non-linear, therefore two models have been applied in analysis of the SSS Study to account for this.

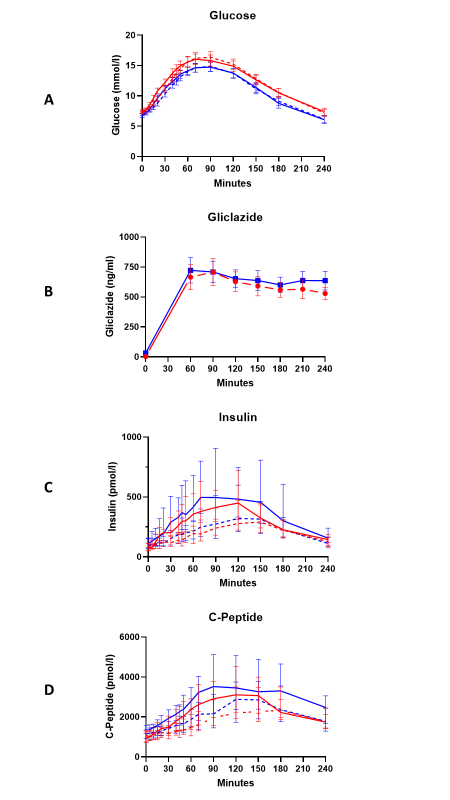

Figure 1: Mean (SEM) plasma glucose (A), gliclazide (B) and Median (IQR) insulin (C) and C-peptide (D) concentrations during OGTT (Solid Lines) and IIGI (Dashed Lines) in Control (Red Lines) and Gliclazide Intervention (Blue Lines)

##### 1.1.2 Parameters of Beta-Cell Modelling

- Parameters of beta-cell modelling will be derived by previously well-defined techniques for assessing insulin secretion by modelling in multiple meal tests (1). The parameters will be analysed by Professor Andrea Mari, of the University of Padua, Italy.
- The model is a mathematical model of the glucose control of insulin secretion capable of quantifying beta-cell function from a physiological meal test.
- Parameters of beta cell modelling were derived in the study of sulphonylurea synergy with incretins as a secondary outcome, which found a 50% augmentation in glucose sensitivity with use of low dose sulphonylurea versus control.

##### 1.1.3 Previous studies utilising beta-cell modelling to compare four intervention groups

- The study by Aaboe et al. (2) was used to provide power to LOGIC Study.
- This study compared the additive effect of GIP infusion or SU alone versus combination on parameters of insulin secretion using intravenous clamps. A similar method will therefore be used to compare the interaction effect between SU/DPP4 versus SU or DPP4 alone.

### 2 Study Objectives and Endpoints

#### 2.1 Study Objectives

| Primary Objective | Outcome Measure | Timepoint of Outcome Measured |
| --- | --- | --- |
| To establish whether a low dose of sulphonylurea will have a synergistic role on glucose sensitivity when given in combination with DPP4 inhibitor | Glucose sensitivity at MMT | Mixed meal tolerance tests at day: 14, 28, 42, 56 |

| Secondary Objective | Outcome Measure | Timepoint of Outcome Measured |
| --- | --- | --- |
| To evaluate the primary outcome for KCNJ11 (rs5219) genotype | Categorical – EE, EK, KK Genotype | Participants will be genotyped at first mixed meal test.  Primary analysis will be repeated adjusting for genotype at baseline covariate |
| To establish the difference in blood glucose with combinations of low dose sulphonylurea and DPP-4 inhibitor | Basal and mean glucose from area under the curve at mixed meal tolerance test  Mean amplitude of glycaemic excursion on continuous glucose monitoring  Time in range on continuous glucose monitoring  Frequency of symptomatic hypoglycaemia | Mixed Meal Tolerance Test days 14, 28, 42, 56  Continuous glucose monitoring during each interventional block |
| To establish pharmacokinetics of low dose gliclazide | Plasma gliclazide levels | Mixed meal tolerance test following low dose sulphonylurea block (SU or SUDPP4)  30 Participants Timepoints - 0 |
| To evaluate whether a low dose of sulphonylurea will have a synergistic role on insulin secretion when given in combination with DPP4 inhibitor | Area Under the Curve INSULIN  Insulin Secretion Rates  Parameters of beta cell modelling  Interaction effect | Mixed meal tolerance tests at day 14, 28, 42, 56 |

### 3 Technical Software

- **Current version of SSS Protoco**l – Version 2.0 27^h^ August 2019
- **Computer Operating System** – Windows 10
- **Data Management Software** – Microsoft Excel for Windows 10
- **Analysis Software –** IBM SPSS Statistics for Windows, Version 26 (3), and R Studio (4).
- **Continuous Glucose Monitoring Software**: Abbott LibreView (5), easyGV (Easy Glycaemic Variability), Oxford University (6)

### 4 Study Methods

#### 4.1 Study Configuration and Experimental Design

- **Research Question**: Does a very low dose of sulphonylurea further augment glucose sensitivity when given in combination with a DPP4 inhibitor?
- **Study Type**: Interventional study
- **Enrolment**: 30 Participants
- **Study Recruitment Sources**:
  - LOGIC Study (NCT03705195),
  - TRIMASTER Study (NCT02653209)
  - SHARE research networks
- **Allocation**: Randomised
- **Interventional Model**: Parallel assignment
- **Clinical Phase Duration**: 8 weeks
- **Interventional Model Description**: Physiological Study
- **Masking**: Open Label
- **Primary Purpose**: Basic Science
- **Controls**: Participants act as own control

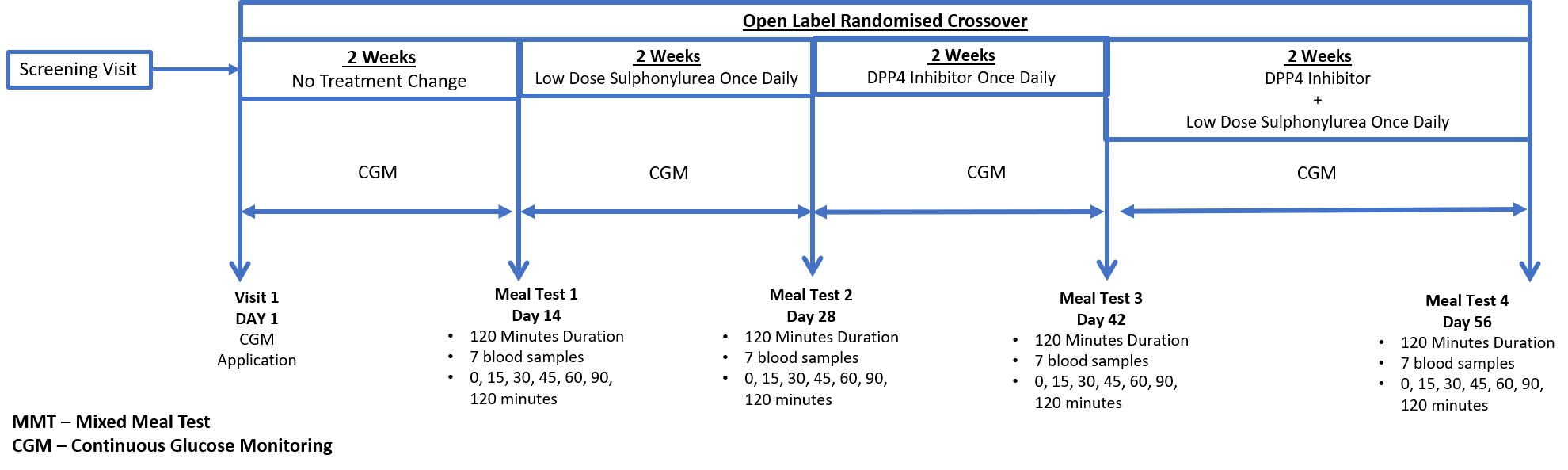

##### 4.1.1 Randomisation

- Each of the 30 participants will complete 4, 2-week blocks during the clinical phase of the study.
- Each participant will complete the blocks in a random order.
- Participants will be randomised using a computer randomisation programme.

| Block | Intervention |
| --- | --- |
| No Intervention | No change to participants standard care |
| Low Dose Sulphonylurea Alone | Gliclazide 20mg Once Daily |
| DPP-4 Inhibitor Alone | Sitagliptin 100mg Once Daily |
| Low dose sulphonylurea + DPP-4 Inhibitor | Gliclazide 20mg Once Daily  +  Sitagliptin 100mg Once Daily |

##### 4.1.2 Meal Tests

- Each participant will complete a meal test at the end of each block i.e. 4 blocks with 4 meal tests in total
- Meal tests will occur on the following days during the 8-week clinical phase
  - Day 14
  - Day 28
  - Day 42
  - Day 56
- Meal test duration is 120 minutes.
  - The participant is given a liquid meal (160mls of Fortisip Compact) at Time 0
  - 7 Blood samples

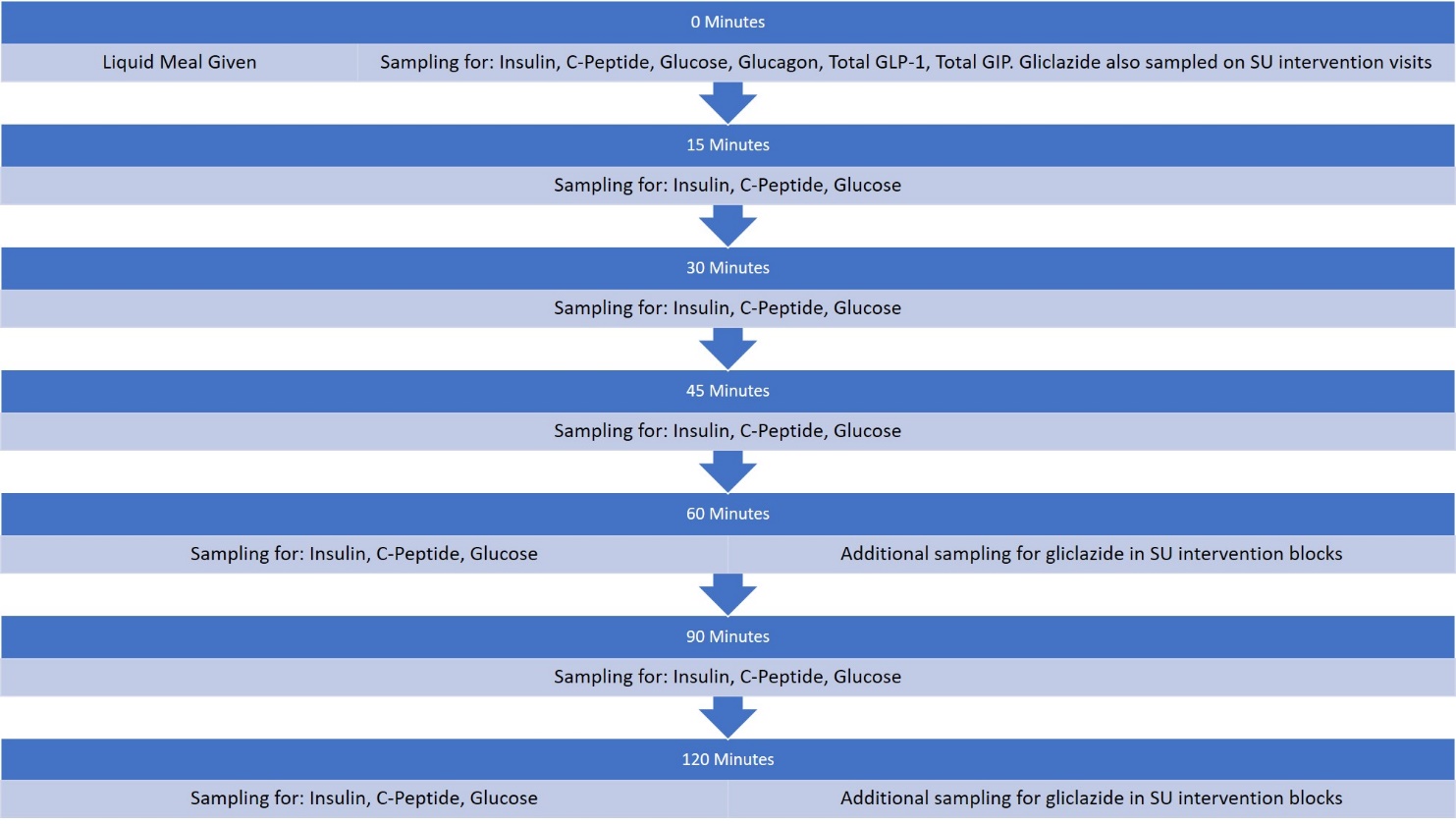

#### 4.2 Eligibility Criteria and Study Population

| Inclusion Criteria | Exclusion Criteria |
| --- | --- |
| - Type 2 Diabetes Mellitus - No treatment or metformin monotherapy - Aged ≥40 and ≤80 years - White British - HbA1c ≤ 8.0% (64mmol/mol) - Estimated Glomerular Filtration Rate ≥ 50ml/min^-1^ - Alanine Transferase ≤ 2.5 * Upper Limit of Normal - Able to Consent | - Type 1 Diabetes Mellitus - HbA1c >8.0% (>64mmol/mol) - Estimated Glomerular Filtration Rate <50ml/min^-1^ - Alanine Transferase > 2.5 * Upper Limit of Normal - Anaemia (Haemoglobin <12.0g/gL for women, <13.0 g/dL for men) - Pregnancy, lactation or a female planning to conceive within the study period - Established pancreatic disease - Participating in clinical phase of another interventional trial/study or have done so within the last 30 days. - Any other significant reason for exclusion as determined by investigator |

Individuals will not be enrolled to the trial/study if they are participating in the clinical phase of another interventional trial/study or have done so within the last 30 days. Individuals who are participating in the follow-up phase of another interventional trial/study, or who are enrolled in an observational study, will be co-enrolled where the CIs of each study agree that it is appropriate.

##### 4.2.1 Number of Participants

- 30 Participants will be recruited in total
- 30 Participants will contribute to the final analysis
- 9 of the 30 Participants will complete the pharmacokinetic phase of study for 24-hour profiling of gliclazide

#### 4.3 Study Measurements

##### 4.3.1 Insulin

- Time Point (Minutes) During Visit: 0, 15, 30, 45, 60, 90, 120
- Day Assessed: Mixed meal tolerance tests on day 14, 28, 42 and 56
- Categorical or Continuous: Continuous

##### 4.3.2 C-Peptide

- Time Point (Minutes) During Visit: 0, 15, 30, 45, 60, 90, 120
- Day Assessed: Mixed meal tolerance tests on day 14, 28, 42 and 56
- Categorical or Continuous: Continuous

##### 4.3.3 Glucose

- Time Point (Minutes) During Visit: 0, 15, 30, 45, 60, 90, 120
- Day Assessed: Mixed meal tolerance tests on day 14, 28, 42 and 56
- Categorical or Continuous: Continuous

##### 4.3.4 Incremental Area Under the Curve

- For insulin, c-peptide and glucose concentrations, incremental values (iAUC, i.e. baseline values subtracted) will be calculated using the trapezoidal rule. AUC will be calculated for each time point as well as total meal test.

##### 4.3.5 Glucagon

- Time Point (Minutes) During Visit: 0
- Day Assessed: Mixed meal tolerance tests on day 14, 28, 42 and 56
- Categorical or Continuous: Continuous

##### 4.3.6 Total GLP-1 and Total GIP

- Time Point (Minutes) During Visit: 0
- Day Assessed: Mixed meal tolerance tests on day 14, 28, 42 and 56
- Categorical or Continuous: Continuous

##### 4.3.7 Gliclazide

- Time Point (Minutes) During Visit: 0, 30, 60, 90, 120 (240, 480, 24 hours for pharmacokinetic participants (n=9))
- Visit assessed: All mixed meal tolerance test visits
- Categorical or continuous: Continuous

##### 4.3.8 KCNJ11 (rs5129) Genotype

- Time Point: Meal test one
- Categorical or continuous: Categorical
- Categorical Outcomes: EE, K Allele
- Analysis to be conducted utilising dominant model (EE vs EK+KK)
- Analysis as baseline covariate as secondary outcome

##### 4.3.9 Parameters of Beta Cell Modelling

##### Parameters of Beta Cell modelling will be derived by Professor Andrea Mari utilising well defined modelling methods (1) (7)

##### Parameters such as Insulin Secretion Rate, Rate Sensitivity, Glucose Potentiation and Potentiation Factor will be derived for each time point.

##### 4.3.10 Parameters from Continuous Glucose Monitoring

###### 4.3.10.1 Estimated HbA1c

- Time Point Measured: derived from continuous glucose monitoring results at day 14, 28, 42, 56
- Categorical or continuous: variable could be either, analysed as continuous for purposes of study
- Parametric or non-parametric: parametric

###### 4.3.10.2 Time in Range

- Time point measured: continuous glucose monitoring during each study block
- Download will produce percentage time in range for three outcomes:
  - Glucose <3.5 mmol/l
  - Glucose 3.5 – 10 mmol/l
  - Glucose >10 mmol/l

###### 4.3.10.3 Mean Amplitude of Glycaemic Excursions (MAGE)

- MAGE represents the mean of blood glucose values exceeding one standard deviation from the 24-hour mean blood glucose and is used as an index of glycaemic variability
- Time Point Measured – continuous glucose monitoring during each study block
- Download will produce average mean glucose values for 2-hour time windows over a 24-hour period from sensor.
- In addition, a daily average glucose will be derived.
- Comparisons will be made between each time window of each treatment group.

###### 4.3.10.4 Glycaemic Risk Assessment Diabetes Equation (GRADE)

- GRADE is a method of quantifying glycaemic profiles into a clinical risk score. It uses all the blood glucose information available to provide a single value defining the assessed clinical risk to which a patient is exposed.
- Non-Diabetic, well controlled glucose profiles yield GRADE scores <5. GRADE scores >5 indicate clinically significant periods of hypo or hyperglycaemia.

###### 4.3.10.5 Mean Glucose

- Parameter of mean glucose for the 14-day sensor

###### 4.3.10.6 Low Blood Glucose Index and High Blood Glucose Index

### 5 Analysis Plan

- As data from LOGIC Study suggest a non-linear response, two modelling methods will be applied.
- Univariate and multivariate descriptive statistics techniques will be used to describe the basic features of the data.

#### 5.1 Level Hierarchy

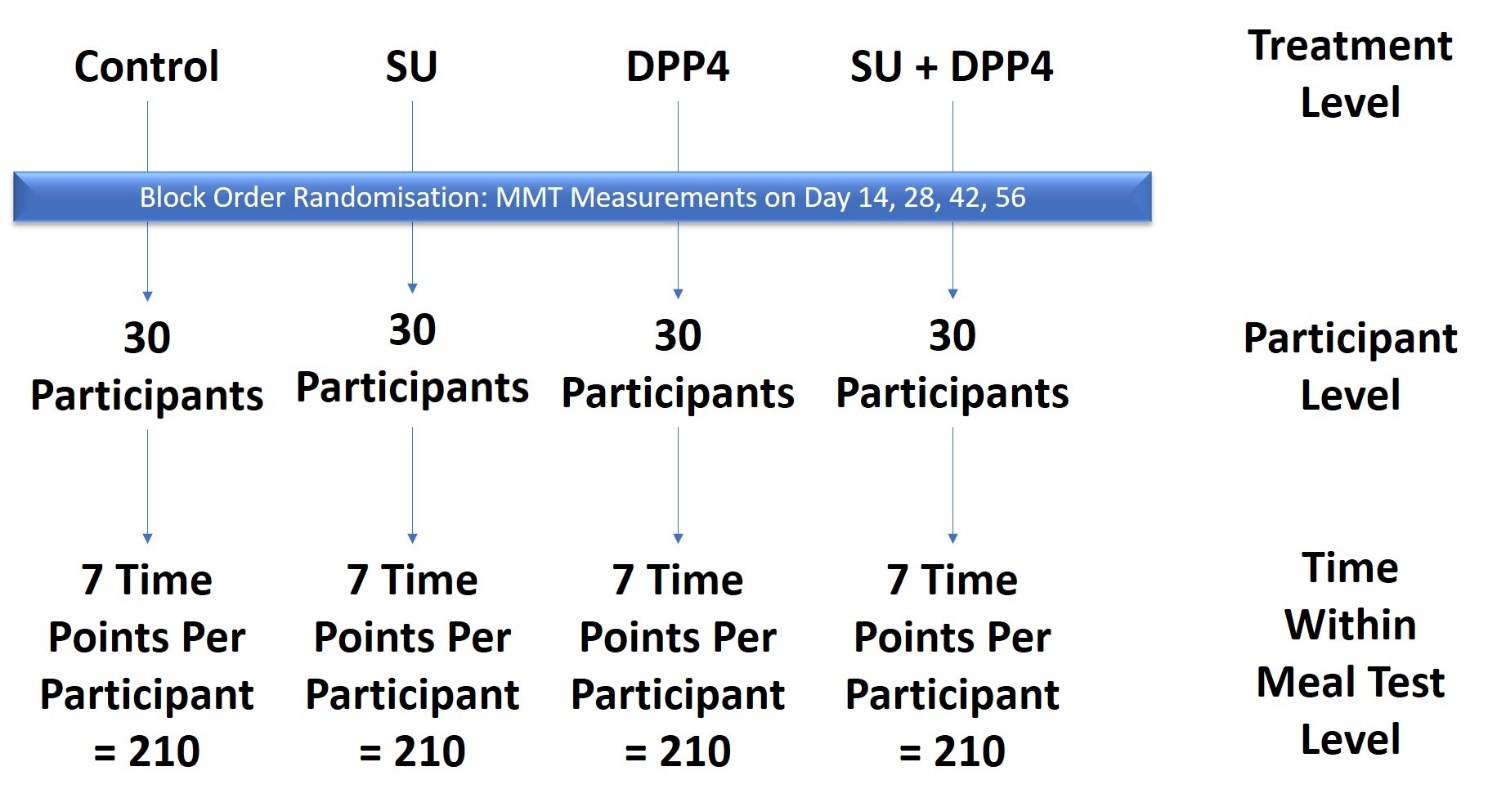

#### 5.2 Linear Mixed Model with Random Effect

- In the first, a linear mixed effects model will be applied using lme4 package in R.
- The dependent variable of glucose sensitivity will be analysed separately for primary outcome:
  - Incremental area under the curve (insulin)
  - Insulin secretion rates.
- The same model will be performed for each measured meal test parameter
- Analysis will be repeated adjusting for baseline covariates including KCNJ11 (rs5219) genotype.
- Treatment level will be determined as a fixed effect
  - There are 4 treatment levels: 1 control and 3 medication intervention
- Time points within the meal test will be fixed or random effect
- “Subject ID”, “Day of MMT” and “Block Number” during clinical phase will be treated as random effects.
- Mixed model will be coded using “lme4” package in R

Formula:

e.g. Y = Glucose Sensitivity

#### Glucose Sensitivity = Treatment + Time point + (1|Subject) + (1|Day) + (1/Block Number) + Ɛ

## Or

**Glucose Sensitivity = Treatment + (1/Time point) + (1|Subject) + (1|Day) + (1/Block Number) + Ɛ**

#### 5.3 Generalized additive mixed models

- As the time course of insulin response in LOGIC study was non-linear, a second analysis applying a generalized additive mixed model will also be conducted.
- In this analysis, “Time” will be considered as a fixed non-linear effect.
- Pharmacokinetic data for gliclazide will also be modelled using a generalized additive mixed model, which will be coded using “mgcv” and “gamm4” packages in R

e.g. Y = Glucose Sensitivity

**Glucose Sensitivity = Treatment +f1(Day)+ f2 (Time point) +(1/Subject) (1/Block number) + Ɛ**

where f1 and f2 are smooth main effects.

### 6 Sample Size

- The primary analysis will be on parameters from the Mixed Meal Tolerance Test.
- Based on previous data in T2DM, the standard deviation of the different in AUC glucose between placebo and vildagliptin treatment was 125 mmol/l 240 minutes (8).
- With 30 participants, the study would have 80% power, p=0.05 to detect a difference of 1/3 of that seen with vildagliptin alone compared with placebo.
- The power would be enough to detect approximately 50% of the different in AUC_INSULIN_:AUC_GLUCOSE_ ratio seen comparing vildagliptin and placebo.

### 7 Timing of Analyses

- Analysis will take place following end of study i.e. last patient last visit.
- Due to COVID-19 pandemic interim analysis for adjustment of sample size will be considered.

### 8 Analysis Populations

##### 8.1.1 Full Analysis Population

- All participants completing SSS Study

##### 8.1.2 Pharmacokinetic Population

- All participants (n=9) completing pharmacokinetic phase of study i.e. 24-hour PK visits during study blocks with low dose gliclazide intervention.

##### 8.1.3 Safety Population

- All subjects participating in SSS STUDY
- All subjects who received any study treatment and are confirmed as providing complete follow-up regarding adverse event information.

#### 8.2 Covariates and Subgroups

- Due to small sample size, power of analysis for covariates and subgroups may be limited.
- Investigators will evaluate baseline data of study prior to completing sub-analysis by subgroup.

| Baseline Co-Variate | Continuous/Categorical |
| --- | --- |
| Sex | Categorical |
| Age | Continuous/Categorical |
| Duration of Diabetes (Years) | Continuous |
| Baseline HbA1c (mmol/mol) | Continuous |
| Height (cm) | Continuous |
| Weight (kg) | Continuous |
| BMI (kg/m2) | Continuous |
| Body Surface Area | Continuous |
| Prior Treatment (Diet/Metformin) | Categorical |

#### 8.3 Missing Data

The research team will make a reasonable attempt to complete all data for each participant. If participants choose to withdraw, the data collected up until the point of withdrawal will be included. If at any point a participant withdraws their consent, all data/samples collected from said individual will be destroyed.

Application of linear and non-linear modelling packages will interpolate data around missing timepoints.

#### 8.4 Interim Analyses and Data Monitoring

- No interim analyses will be conducted originally planned for study.
- Amendments to study following interim analysis will be preserved.

### 9 Summary of Study Data

#### 9.1 Datasets

##### 9.1.1 Master Parameters Information: Definitions and variable codes

##### 9.1.2 Baseline Dataset: Includes baseline covariate information for all participants

##### 9.1.3 Master Dataset: Long format data matrix including all study parameters for use in main analysis. Columns will be added for beta cell modelling parameters

##### 9.1.4 Pharmacokinetic Dataset: Participants SSS1 – SSS9 only for analysis of gliclazide pharmacokinetics

##### 9.1.5 Incretin Dataset: Measurements of incretin hormones, time point 0 only

#### 9.2 Null Hypothesis

- The null hypothesis is that the Primary Outcome is not different from zero

#### 9.3 Secondary Analysis for KCNJ11 Genotype

- Dominant model will be utilised for analysis by KCNJ11 genotype.
- Primary analysis will be repeated for EE homozygotes and those possessing the K-allele (EK +KK).

#### 9.4 Presentation of Data

- Parameters of insulin, c-peptide, glucagon, GLP-1 and GIP are not predicted to follow Gaussian distribution; therefore, data will be presented as median (lower quartile, upper quartile).
- As glucose and gliclazide data are predicted to follow Gaussian distribution, data will be presented as mean ± standard error of mean
- Parameters of beta-cell modelling will be presented as mean ± standard error of mean

#### 9.5 Subject Disposition

- All participants who were recruited completed SSS study within the required timeframe.

#### 9.6 Demographic and Baseline Variables

Summary statistics will be produced in accordance with section 8.

#### 9.7 Concurrent Illnesses and Medical Conditions

Concurrent illnesses and medications will be coded according to ICD 10.

#### 9.8  Prior and Concurrent Medications

Only concurrent and new medications will be documented as part of SSS study and coded as per the British National Formulary.

#### 9.9 Treatment Compliance

SSS study medication compliance is assessed during telephone calls. Compliance will be assessed by tablet number returned at meal test visits. If a participant is unable to be compliant, they will be excluded from study. An accountability log will be kept as per TASC SOP37.

### 10 Safety Analyses

#### 10.1 Adverse Events

- The summary statistics of adverse events will be produced at end of study.

#### 10.2 Deaths, Serious Adverse Events and other Significant Adverse Events

- Summary statistics will be produced in accordance with sponsor guidance.
- Data will be presented as median (IQR)

#### 10.3  Pregnancies

Pregnancy tests will be performed on all women of child-bearing age at study screening prior to recruitment.

#### 10.4  Clinical Laboratory Evaluations

The summary statistics will be produced in accordance with section ‎0.

#### 10.5 Other Safety Measures

‘The summary statistics will be produced in accordance with section ‎0.’ Vital signs might be appropriately included in this subsection. Many of the points made regarding laboratory tests in section ‎13.4 are relevant to vital signs.

### 11  Pharmacokinetics

Gliclazide pharmacokinetics will be analysed as part of SSS study. Analysis will include gliclazide concentration over time, calculation of C-max, T-max, half-life and time to steady state. Further sub-analysis of gliclazide concentration will be made accounting for co-variates outlined in section 9.3 in addition to KCNJ11 genotype.

### 12 Other Analyses

Modelling of beta cell function will be conducted by Professor Andrea Mari of The University of Padua, Italy as per previously well reported methods (1, 7, 9, 10).

### 13 Remit of the Report document

The statistical analysis section for final report will be extracted from SAP.

### 14  Reporting Conventions

- P-values ≥0.001 will be reported to 2 decimal places; p-values less than 0.001 will be reported as “<0.001”.
- Estimated parameters, not on the same scale as raw observations (e.g. regression co-efficient) will be reported to 3 significant figures.

3.	IBM Corporation A, New York, USA. IBM SPSS Statistics for Windows 2020.

4.	R: A language and environment for statistical computing. . Vienna, Austria2014.

5.	Care AD. LibreView for Freestyle Libre Pro Version 1.0. USA2018.

6.	Oxford Uo. EasyGV. In: Hill N, editor. Diabetes Technology and Therapeutics2011.

7.	Mari A, Schmitz O, Gastaldelli A, Oestergaard T, Nyholm B, Ferrannini E. Meal and oral glucose tests for assessment of beta -cell function: modeling analysis in normal subjects. Am J Physiol Endocrinol Metab. 2002;283(6):E1159-66.

8.	Ahren B, Pacini G, Foley JE, Schweizer A. Improved Meal-Related  -Cell Function and Insulin Sensitivity by the Dipeptidyl Peptidase-IV Inhibitor Vildagliptin in Metformin-Treated Patients With Type 2 Diabetes Over 1Year. Diabetes Care. 2005;28(8):1936-40.

9.	Mari A, Ferrannini E. Beta-cell function assessment from modelling of oral tests: an effective approach. Diabetes Obes Metab. 2008;10 Suppl 4:77-87.

10.	Mari A, Pacini G, Murphy E, Ludvik B, Nolan JJ. A Model-Based Method for Assessing Insulin Sensitivity From the Oral Glucose Tolerance Test. Diabetes Care. 2001;24(3):539-48.
